# Statin Exposure and Risk of Dialysis in Type 2 Diabetes: A Real-World Cohort Study

**DOI:** 10.64898/2026.05.14.26353258

**Authors:** César Augusto Madid Truyts, Amanda Gomes Rabelo, Maria Tereza Abrahao, Mateus de Lima Freitas, Edson Amaro Junior, Rogério da Hora Passos, Adriano José Pereira

## Abstract

1

**Background:** Renal effects of statins in type 2 diabetes mellitus (T2DM) remain uncertain. We evaluated whether statin exposure is associated with time to dialysis initiation.

**Methods:** We conducted a retrospective cohort study of adults with T2DM, indexing follow-up at diagnosis during first hospital admission (day 0) between january 2017 and march 2025. Statin use was modeled as time-varying from statin days; (classified in 3 categories: baseline users, new users, and never users). The primary outcome was dialysis. Analysis estimated cause-specific hazards, censoring deaths; proportional hazards were checked with prespecified windows of statin exposure (0–1, 1–3, > 3 years). Competing-risk analyses (Fine–Gray) assessed the sub-distribution hazard of dialysis with death as a competing event in two models: (i) prevalent users at baseline and (ii) new-users with post-initiation intervals of 30 and 90 days. An Observational Medical Outcomes Partnership Common Data Model standardized dataset of a Brazilian quaternary hospital, and the Real-World Data tool MD Clone were used in the study.

**Results:** Of 36,246 adults identified, 32,125 entered the time-varying cohort (39,943 risk intervals; 656 dialysis events); median follow-up among censored patients was 753 days. At baseline, 70.3% never used statins, 5.5% were users (≤ 0 days), and 24.2% initiated after diagnosis. Crude dialysis incidence was 4.51 vs. 12.31 per 1,000 patient-years during unexposed vs. exposed time. In the adjusted time-varying Cox model, current statin exposure was associated with a modestly higher hazard of dialysis (HR = 1.043, 95% CI 1.011–1.077). In the new-users analysis, HRs were 0.83 (95% CI 0.66–1.05), and 0.73 (95% CI 0.57–0.92) with a 30-day and 90-day intervals, respectively.

**Conclusions:** In this retrospective cohort of hospitalized diabetic patients at baseline, statin initiation at least 90-days in advance is associated with reduced indication of renal replacement therapy.

## 2 Introduction

Type 2 diabetes mellitus (T2DM) is the leading global cause of chronic kidney disease (CKD) and end-stage kidney disease [1]. Lipid-lowering therapy is widely prescribed to reduce atherosclerotic cardiovascular risk, and contemporary guidance recommends statin or statin/ezetimibe for adults with reduced estimated glomerular filtration rate (eGFR) [2]. While cardiovascular benefits are well established, the renal effects are modest and heterogeneous. Statins may reduce proteinuria or slow the decline in eGFR; however, consistent reductions in kidney failure events have not been demonstrated [3, 4]. In patients with T2DM, evidence on renal effects after statin initiation is heterogeneous: RCTs and meta-analyses report modest molecule-specific improvements in albuminuria or eGFR, whereas extensive observational data show variable risks across kidney outcomes [5–8].

Despite extensive prescribing and accumulating safety data, a key knowledge gap persists whether statin exposure among patients with T2DM is associated with increased need of renal replacemente therapy (RRT). Previous studies present limitations when keeping together acute and chronic renal outcomes [9], applying time-fixed exposure definitions (introducing “immortal-time” and prevalent-user biases) [10–12], and when rarely model treatment as time-varying or account for the competing risk of death[13–16]. We therefore evaluated the association between time-updated statin exposure and time-to-dialysis in a real-world cohort of T2DM, estimating cause-specific hazards of dialysis initiation.

## 3 Methods

### 3.1 Study design and reporting

The reporting of this retrospective cohort study followed the Strengthening the Reporting of Observational Studies in Epidemiology (STROBE) guideline. This study was approved by the Research Ethics Committee of Einstein Hospital Israelita, (CAAE 79489824.0.0000.0071, July 3rd, 2024). Given its retrospective design, which utilizes de-identified electronic health record (EHR) data, the requirement for informed consent was waived. All procedures complied with the Declaration of Helsinki, applicable Brazilian regulations, Brazilian Data Protection Legislation.

### 3.2 Setting and data source

This study was conducted using EHR data from Einstein Hospital Israelita, a quaternary care center in São Paulo, Brazil. Data were accessed through the MDClone ADAMS platform (version 11.1., Beer-Sheva, Israel), which queries an underlying database previously mapped to the Observational Medical Outcomes Partnership Common Data Model [17].

### 3.3 Participants

We included adults (≥18 years) with T2DM identified from diagnosis codes recorded in the institutional database (Supplementary Table S1). The index date (day 0) was defined as the date of first hospital admission with T2DM between january 2017 and march 2025.

To reduce misclassification, patients with codes compatible with type 1 diabetes mellitus (ICD-10 E10.x and O24.0) were excluded. Given the variability of coding in routine clinical practice, the remaining cohort included patients with type 2 diabetes mellitus (E11.x), unspecified diabetes (E14.x), and other non–type 1 diabetes codes (e.g., E13.x and E12.x), as well as diabetes-related complications recorded without a concurrent etiologic diabetes code.

Because diabetes-related complications (e.g., diabetic retinopathy, neuropathy, or nephropathy) may be recorded without an explicit diabetes diagnosis code in EHR, such codes were also considered indicative of underlying diabetes.

This approach reflects real-world documentation patterns and prioritizes sensitivity in identifying adults with non–type 1 diabetes in routine clinical care.

Patients were followed longitudinally after discharge using outpatient and subsequent inpatient records. Follow-up continued until the earliest occurrence of dialysis initiation, death, or last available clinical record.

### 3.4 Exposure definition

Statin exposure was modeled as a time-varying variable based on statin prescription records. Patients contributed person-time as unexposed from index date until statin initiation and as exposed thereafter.

For secondary analyses, exposure was parameterized using: (i) baseline statin use (prevalent users, defined as statin use on or before index date), and (ii) a new-user design, comparing initiators (statin initiation after index) with never users.

In the new-user analyses, post-initiation lag periods (grace periods) of 30 and 90 days were applied. Among initiators, events occurring within the lag period were censored at the corresponding lag time to mitigate protopathic bias.

### 3.5 Outcome definition

The primary outcome was initiation of RRT. Event dates were obtained from procedure and clinical records.

### 3.6 Covariates

Baseline covariates included age, sex, body mass index (BMI), glycated hemoglobin (HbA1c), serum creatinine closest to the index date, hypertension, cardiovascular or cerebrovascular disease (CVD), and use of angiotensin-converting enzyme inhibitors (ACEi) or angiotensin receptor blockers (ARB).

Continuous covariates were winsorized at the 0.5th and 99.5th percentiles and standardized using median and median absolute deviation (MAD).

### 3.7 Statistical analysis

Baseline characteristics were summarized using medians (interquartile range, IQR) for continuous variables and counts (percentages) for categorical variables.

Time-to-event analyses were conducted using Cox proportional hazards models with time-varying exposure in a counting-process framework. Cause-specific hazards for dialysis were estimated treating death as a censoring event. Models were fitted using L2-penalized (ridge) partial likelihood and reported as hazard ratios (HRs) with 95% confidence intervals (CIs).

Proportional hazards assumptions were assessed using a prespecified piecewise approach (0–1, 1–3, and >3 years since index).

To account for competing risk of death, Fine–Gray subdistribution hazard models were fitted to estimate the cumulative incidence of dialysis.

For descriptive analyses, cumulative incidence functions (CIFs) were estimated using the Aalen–Johansen estimator and compared between groups using Gray’s test. The CIF represents the cumulative probability of experiencing the event of interest over time in the presence of competing risks.

Crude incidence rates were calculated per 1,000 person-years according to time-updated exposure status.

All statistical tests were two-sided with a significance level of *α* = 0.05. Analyses were performed using Python (pandas, lifelines, matplotlib, version 3.11.6, Python Software Foundation, Wilmington, DE, USA)) and R (cmprsk, version 4.5.1, R Foundation for Statistical Computing, Vienna, Austria).

## 4 Results

We identified 36,246 adults with T2DM. After excluding records with invalid or absent follow-up, 32,125 patients were included in the time-varying cohort, contributing 39,943 risk intervals and 656 dialysis events; among censored individuals, the median follow-up was 753 days.

Exposure profile and crude incidence. At baseline, 25,474 (70.3%) had never used statins, 2,004 (5.5%) were already on statins (≤ 0 days), and 8,768 (24.2%) initiated after index (> 0 days). In time-updated person-time, the crude incidence of dialysis was 4.51 and 12.31 per 1,000 person-years during unexposed and exposed periods, respectively.

Table 1 summarizes baseline characteristics and unadjusted group comparisons (statin vs. no statin). Cause-specific hazards - In the cause-specific Cox model adjusted for age, sex, BMI, HbA1c, baseline creatinine, hypertension, CVD, and ACEi/ARB use, current statin exposure was associated with a modestly higher hazard of initiating RRT (HR=1.04, 95% CI 1.01–1.08; *p* = 0.01) (Table 2. Higher baseline creatinine (per robust SD; *p* < 0.01) was strong independent predictors of RRT. Effects of age, sex, BMI, HbA1c, CVD, and ACEi/ARB were small and not consistently significant. Simon-Makuch curves by time-varying statin use showed a slightly higher cumulative incidence among exposed patients, consistent with the Cox estimate (Figure 1).

**Table 1:**
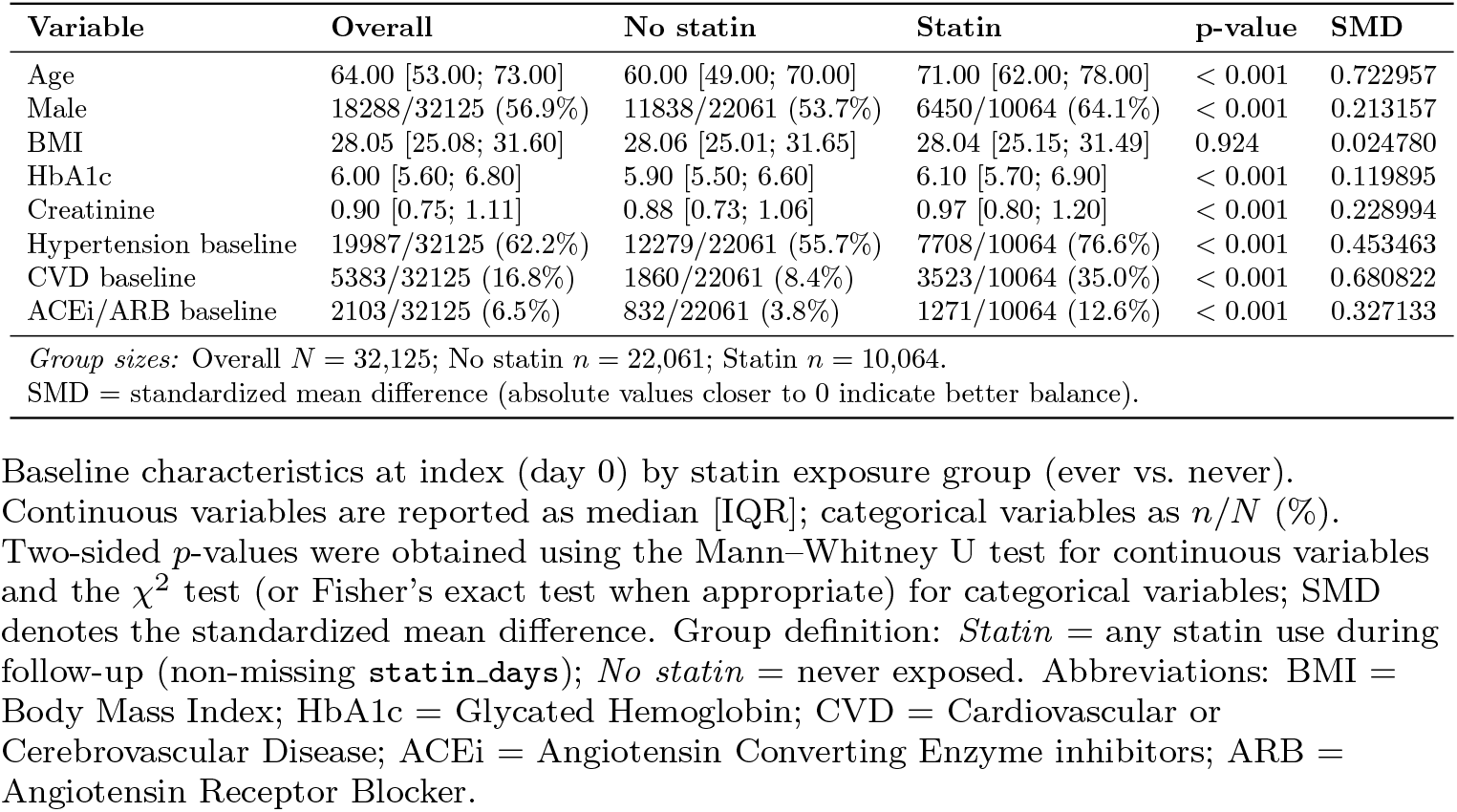
Baseline characteristics by statin exposure group (ever vs. never). Medians [IQR] for continuous variables and *n/N* (%) for categorical variables.

**Table 2:** Time-varying Cox model for time to dialysis initiation in adults with T2DM.

| Covariate | HR | 95% CI | p-value |
| --- | --- | --- | --- |
| Statin exposure (time-updated) | 1.04 | 1.01–1.08 | 0.01 |
| Age (per robust SD)<sup>a</sup> | 1.00 | 1.00–1.00 | 0.05 |
| Male sex | 1.02 | 0.99–1.05 | 0.22 |
| BMI (per robust SD)<sup>a</sup> | 1.00 | 1.00–1.00 | 0.51 |
| HbA1c (per robust SD)<sup>a</sup> | 1.00 | 0.99–1.02 | 0.46 |
| Serum creatinine (per robust SD)<sup>a</sup> | 1.17 | 1.15–1.20 | < 0.01 |
| Hypertension at baseline | 1.02 | 0.99–1.05 | 0.11 |
| Cardio/cerebrovascular disease at baseline | 1.04 | 1.00–1.07 | 0.04 |
| ACEi/ARB use at baseline | 1.02 | 0.96–1.08 | 0.55 |
| *Model summary* |  |  |  |
| Subjects = 32,125; Periods = 39,943; Events = 656 |  |  |  |
| Partial log-likelihood = [update after rerunning model]; Partial AIC = [update] |  |  |  |
| Likelihood-ratio test = [update] on 9 df ($p$ = [update]) | | | |

Cause-specific hazard estimated with a Cox proportional hazards model with time-varying statin exposure (counting-process specification), fitted via L2-penalized partial likelihood (penalizer = 0.5). Death before dialysis was treated as censoring. HR = hazard ratio; CI = confidence interval; BMI = body mass index; ACEi = angiotensin-converting enzyme inhibitor; ARB = angiotensin receptor blocker.
<sup>a</sup> Continuous covariates were winsorized (0.5–99.5%) and standardized using median/MAD; HRs are per robust standard deviation.

**Figure 1:**
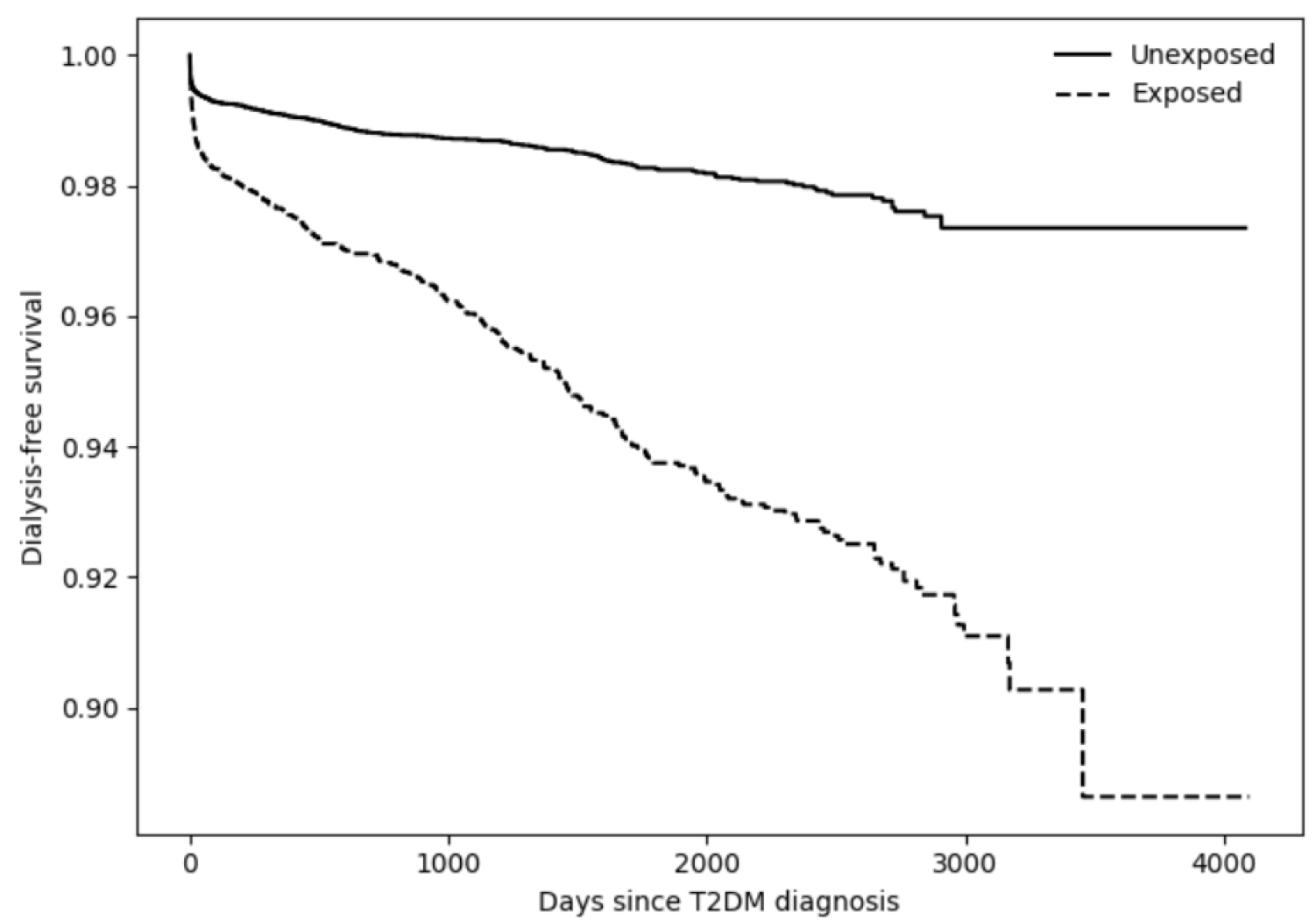
Simon–Makuch dialysis-free survival by time-updated statin exposure. Curves were estimated using left-truncated Kaplan–Meier fits on counting-process intervals [*start, stop*), switching individuals from the unexposed to the exposed curve at statin initiation. Death prior to dialysis was treated as censoring (cause-specific hazard framework). Shaded bands denote 95% confidence intervals; widening at late times reflects small numbers at risk. The exposed curve lies below the unexposed curve, consistent with the adjusted Cox estimate (HR ≈ 1.04).

Proportional-hazards check (piecewise). As shown in Table 3, window-specific statin effects were near null and non-significant across all periods—0–1 year HR=1.02 (0.99-1.06), 1–3 years HR=1.01 (0.98-1.05), and > 3 years HR=1.03 (0.99-1.07), all *p* > 0.20 arguing against a time-restricted excess risk. Baseline creatinine remained significant predictors.

**Table 3:** Piecewise time-varying Cox model for time to dialysis initiation in adults with T2DM.

| Covariate | HR | 95% CI | p-value |
| --- | --- | --- | --- |
| Statin exposure, 0–1 year | 1.02 | 0.99–1.06 | 0.25 |
| Statin exposure, 1–3 years | 1.01 | 0.98–1.05 | 0.48 |
| Statin exposure, >3 years | 1.03 | 0.99–1.07 | 0.21 |
| Age (per robust SD)<sup>a</sup> | 1.01 | 1.00–1.02 | 0.16 |
| Male sex | 1.01 | 0.99–1.03 | 0.37 |
| BMI (per robust SD)<sup>a</sup> | 1.00 | 1.00–1.00 | 0.62 |
| HbA1c (per robust SD)<sup>a</sup> | 1.00 | 1.00–1.00 | 0.58 |
| Serum creatinine (per robust SD)<sup>a</sup> | 1.03 | 1.02–1.03 | < 0.01 |
| Hypertension at baseline | 1.01 | 0.99–1.03 | 0.24 |
| Cardio/cerebrovascular disease at baseline | 1.02 | 0.99–1.05 | 0.13 |
| ACEi/ARB use at baseline | 1.01 | 0.97–1.05 | 0.64 |
| *Model summary* |  |  |  |
| Subjects = 32,125; Periods = 77,793; Events = 656 |  |  |  |
| Partial log-likelihood = [update after rerunning model]; Partial AIC = [update] |  |  |  |
| Likelihood-ratio test = [update] on 11 df ($p$ = [update]) | | | |

Cox proportional hazards model in counting-process form with time-varying exposure parameterized piecewise (0–1, 1–3, and >3 years). Death prior to dialysis was treated as censoring (cause-specific hazard). HR = hazard ratio; CI = confidence interval; BMI = body mass index; ACEi = angiotensin-converting enzyme inhibitor; ARB = angiotensin receptor blocker.
<sup>a</sup> Continuous covariates were winsorized (0.5–99.5%) and standardized using median/MAD; HRs are per robust standard deviation.
Note: For the piecewise model, robust standardization was recomputed on the interval-expanded dataset; HRs per robust SD are therefore not directly comparable to those from the primary model. Interpret within-model.

Treating death as a competing event, baseline statin use (≤ 0 days) was not associated with a lower subdistribution hazard of dialysis after multivariable adjustment (HR = 0.80, 95% CI 0.55–1.15; *p* = 0.220; Figure 2). By contrast, unadjusted CIFs were higher among baseline statin users early after diagnosis (e.g., at 1,000 days: 2.77% vs. 1.72%; Gray’s test *p* = 0.006), and the curves later cross (Figure 3). This prevalent-user contrast targets a different estimand than the cause-specific, time-varying analysis of current use (HR ≈ 1.04).

**Figure 2:**
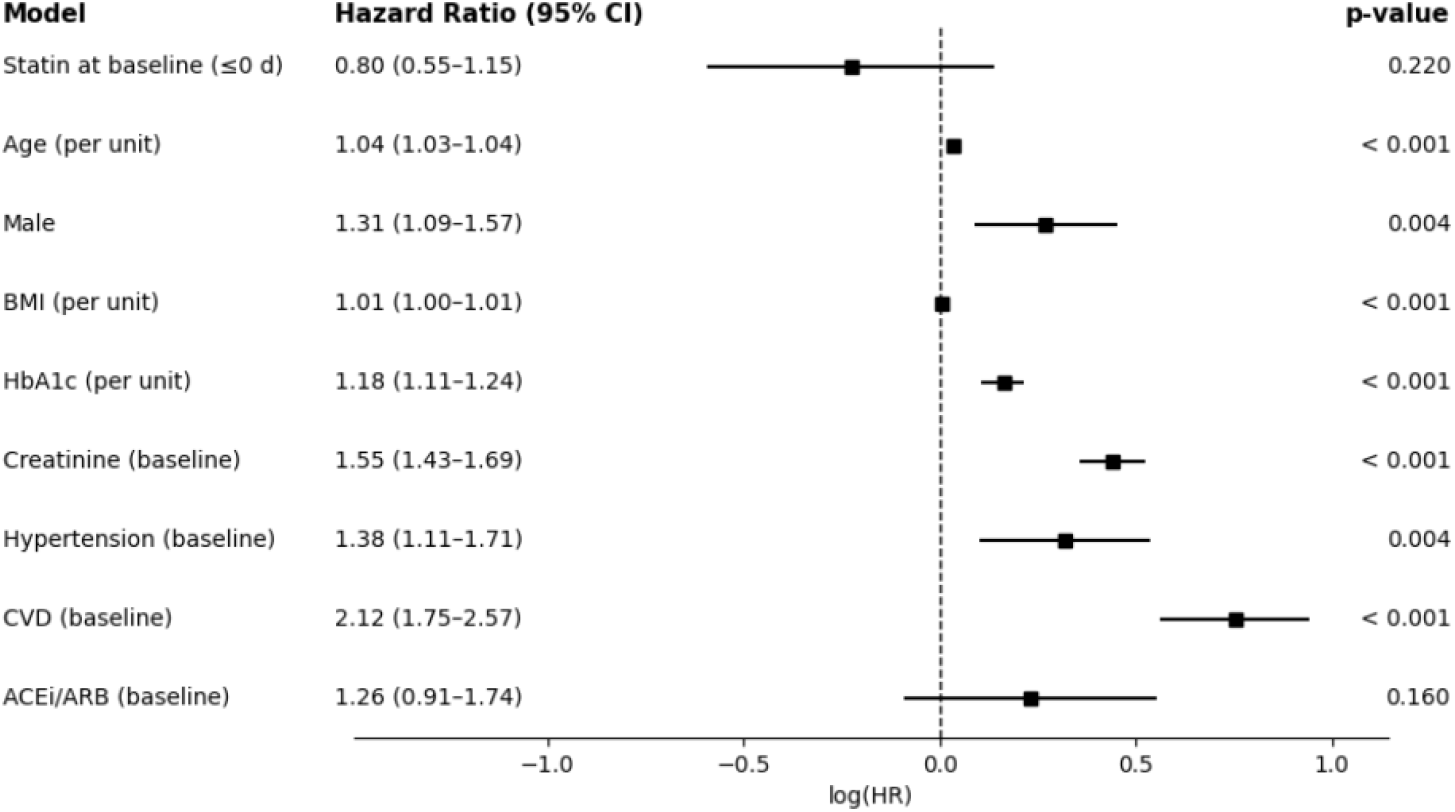
Forest plot of subdistribution hazard ratios (HR) for initiation of dialysis with death as a competing event, estimated with a Fine–Gray model in adults with T2DM. Squares show point estimates and horizontal lines the 95% CIs on the log(HR) scale; the dashed vertical line marks no effect (HR=1), with values to the left indicating a lower subdistribution hazard. “Statin at baseline” denotes prevalent users at index (statin days ≤ 0). The model adjusts for age, sex, BMI, HbA1c, baseline creatinine, hypertension, CVD, and ACEi/ARB use. Abbreviations: BMI = body mass index; HbA1c = glycated hemoglobin; CVD = cardiovascular/cerebrovascular disease; ACEi/ARB = angiotensin-converting enzyme inhibitors/angiotensin receptor blockers.

**Figure 3:**
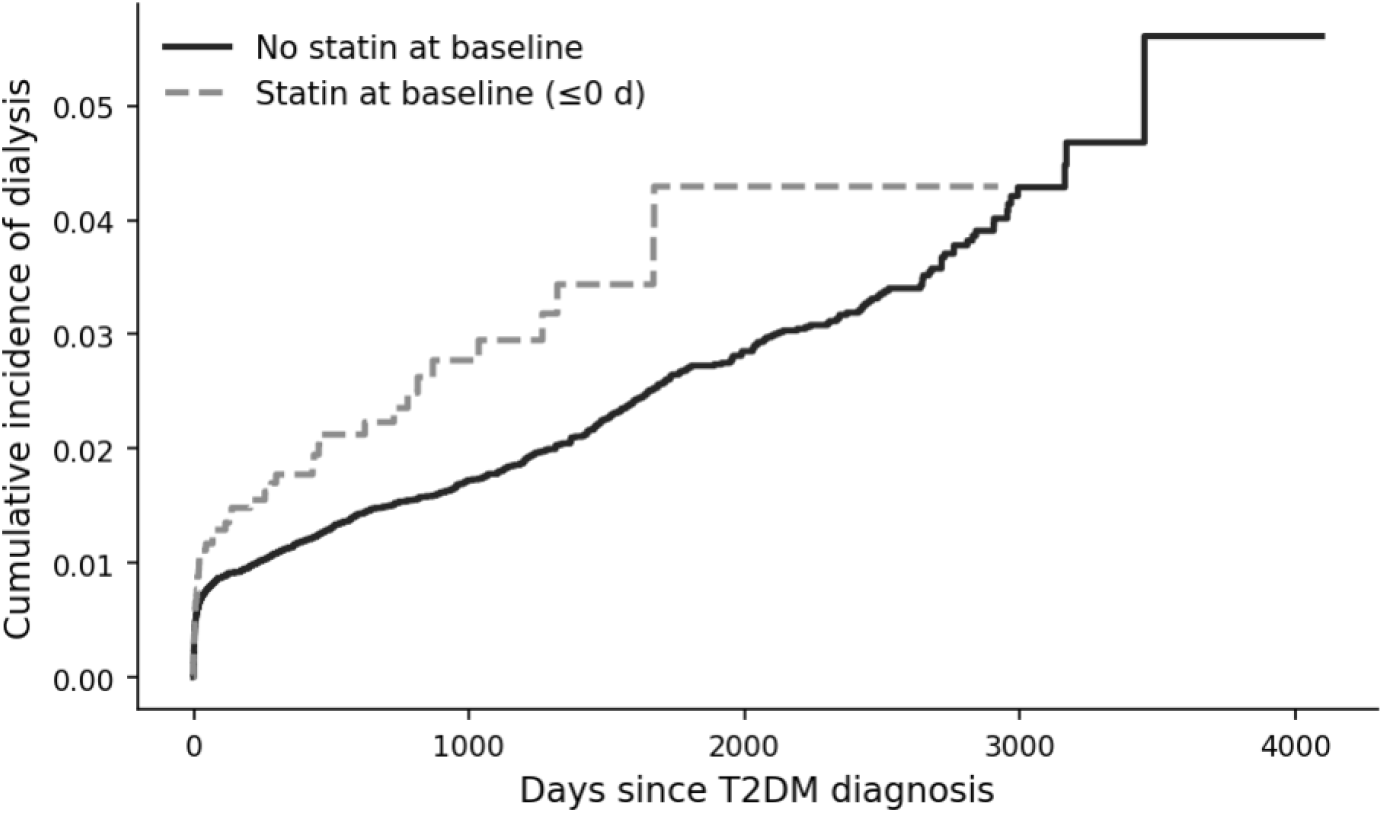
Cumulative incidence function (CIF) for initiation of dialysis with death as a competing event (Aalen–Johansen), stratified by baseline statin use. Solid line: no statin at baseline; dashed line: statin at baseline (≤ 0 d). Example at 1,000 days: 2.77% vs. 1.72%; Gray’s test *p* = 0.006.

In models restricted to new users and applying post-initiation lags, the subdistribution hazard was HR=0.81 (95% CI 0.64–1.04; *p* = 0.10) with a 30-day lag and HR=0.73 (95% CI 0.57–0.93; *p* = 0.01) with a 90-day lag (Figure 4). Unadjusted CIFs were higher among initiators but attenuated with longer lags (significant Gray tests), being consistent with potential confounders (Figure 5).

**Figure 4:**
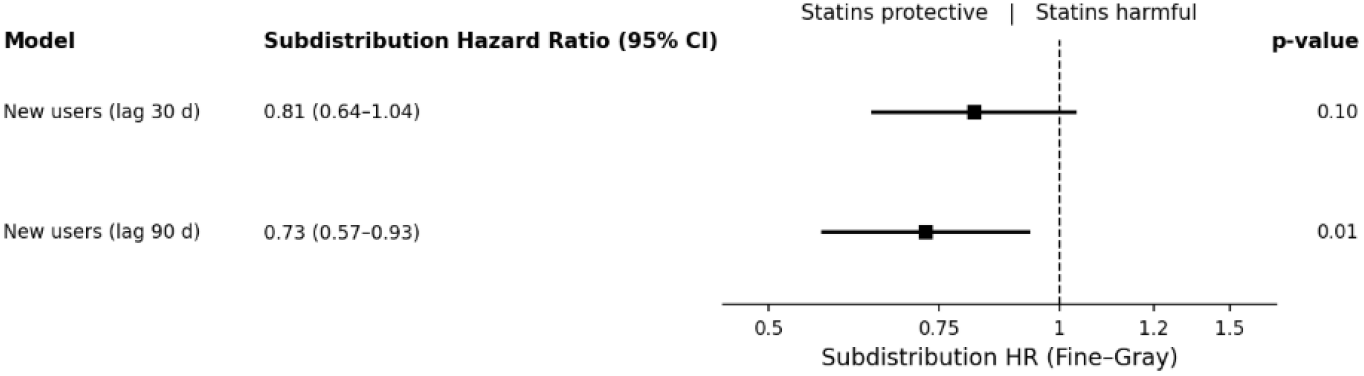
Fine–Gray subdistribution hazard for dialysis (death as competing risk) in a new-user design with post-initiation lags (30 and 90 days). Squares show HR and horizontal lines the 95% CIs on the log(HR) scale; the dashed line marks no effect (HR=1).

**Figure 5:**
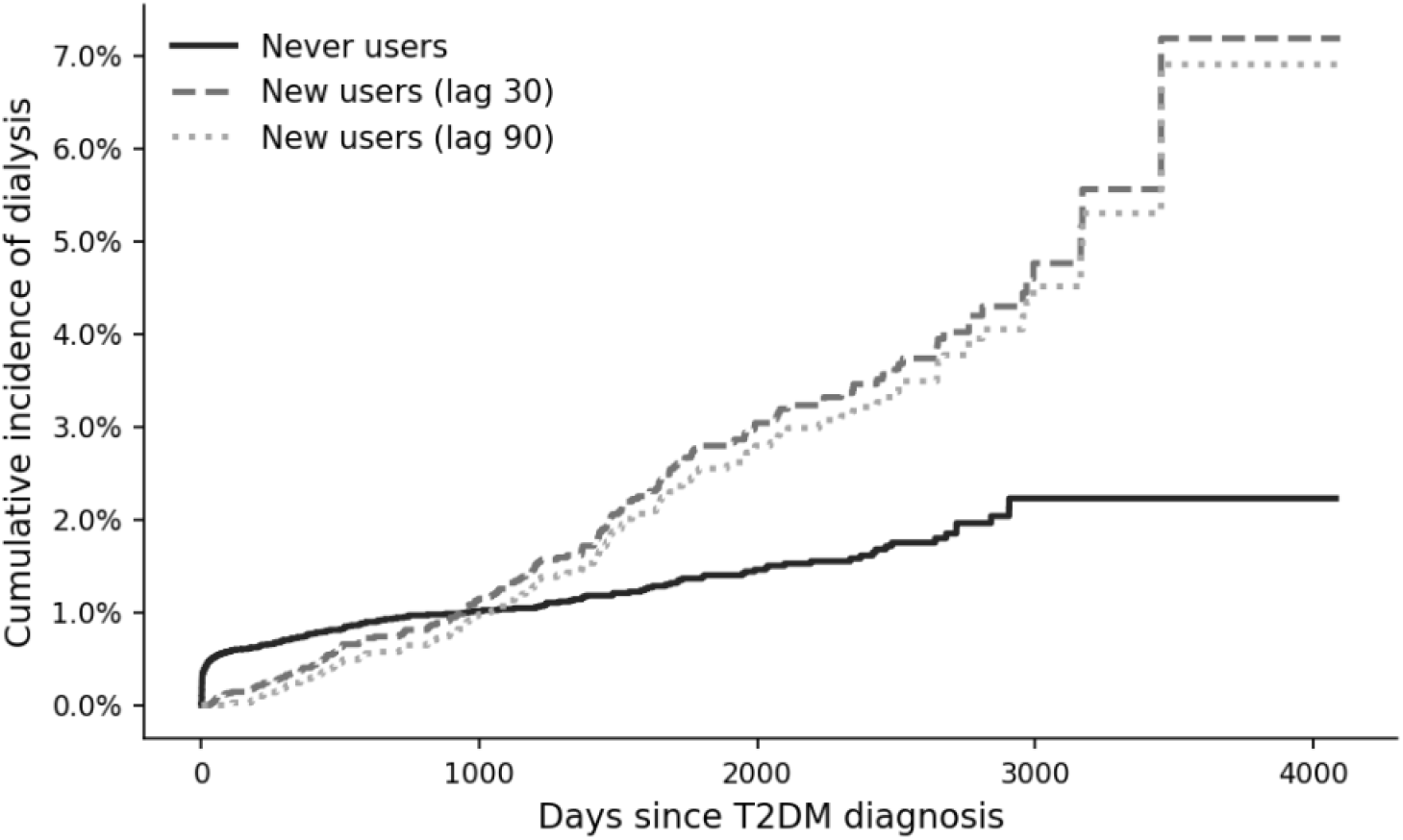
Cumulative incidence of dialysis with death as a competing event (Aalen– Johansen) in a new-user design with post-initiation lags. Curves compare never users (solid) with statin initiators under 30-day (dashed) and 90-day (dotted) lags. CIFs are unadjusted; the lag censors events occurring within the grace period after initiation, mitigating protopathic bias (early incidence is lower with the 90-day lag).

## 5 Discussion

In this real-world T2DM cohort, we found no evidence that statins delay progression to dialysis when modeling current exposure in a cause-specific framework, conversely, the primary time-varying Cox model suggested a small adverse association of about 4% increase in the need of dialysis ((HR ≈ 1.04), CI 1.01-1.08), and prespecified window-specific estimates (0–1, 1–3, > 3 years) were near null, arguing against a time-restricted effect. Simon-Makuch curves likewise showed only modest separation between exposed and unexposed trajectories.

Findings from the competing-risk analyses highlight that different estimands can yield different conclusions. Treating death as a competing event, baseline statin use was associated with a lower sub-distribution hazard of dialysis after adjustment (Fine-Gray HR ≈ 0.80, CI 0.55-1.15). Among new-users, applying post-initiation lags to mitigate protopathic bias, the sub-distribution hazard was not statistically significant for a 30-day lag,(0.81, (95% CI 0.64–1.04), suggesting a modest protective association on the cumulative incidence scale once very early post-initiation events are down-weighted. Taken together, the cause-specific results (instantaneous risk among those still at risk) and the subdistribution results (cumulative incidence under competing death) are not contradictory; they target different quantities and, jointly, do not support a large renal harm signal while leaving room for a modest reduction in dialysis incidence under new-user, lagged specifications.

Prior literature is mixed and often targets different outcomes. In non-dialysis CKD, a recent network meta-analysis reported that rosuvastatin and atorvastatin reduced major adverse cardiovascular events and modestly increased mean eGFR versus control, but dialysis was not a primary endpoint and small eGFR gains may not translate into fewer kidney failure events [18]. Among T2DM specifically, a large observational study found lower risk of incident kidney disease after statin initiation, particularly with intensive LDL-C control [8]; differences in outcome definition (incident CKD vs. dialysis), intensity of risk-factor management, and residual confounding likely contribute to divergent conclusions relative to our dialysis-focused analysis.

Safety considerations appear to be agent/dose specific. Contemporary data in older adults show no material change in kidney function attributable to statins, regardless of baseline CKD, supporting decisions motivated primarily by cardiovascular risk in this age group [19]. Conversely, rosuvastatin has been associated with higher risks of hematuria and proteinuria in observational cohorts [20], and rare cases of reversible tubular toxicity have been reported, particularly at high dose [21]. We could not examine molecule/dose specific effects, which limit safety inferences in this context.

This study has limitations. Despite modeling statin use as a time-varying exposure in counting-process form and adjusting for multiple covariates, residual confounding and protopathic bias remain possible. The new-user analyses with 30–90-day lags mitigate, but do not eliminate—the latter. Small number of patients with baseline laboratory tests available, and the unavialbility of albuminuria and lipid levels (LDL-C) to bettter fit the models may have limited the control for covariates. Exposure misclassification is also possible given unmeasured adherence, temporary discontinuation, and the lack of information on statin class and dose. Outcome misclassification (acute kidney injury requiring dialysis vs. progression to end-stage kidney disease) and uncertainty in event dates cannot be fully excluded. Finally, generalizability may be constrained to similar settings of this real-world T2DM cohort (quaternary hospital, in a large city of a low-middle-income country).

Our findings suggest that statin therapy in T2DM should not be expected to prevent dialysis on etiologic (cause-specific) grounds (it may even worsen it). Its established role remains related to cardiovascular risk reduction. Nonetheless, after mitigating protopathy in a new-user design, we observed a modest reduction in the cumulative incidence of dialysis, a signal that warrants confirmation in prospective trials. Future work should also evaluate heterogeneity by baseline eGFR and albuminuria, compare statin classes and intensities, incorporate lipid targets, and consider active-comparator designs with propensity-based adjustment.

## Data Availability

The datasets generated and/or analyzed during the current study are not publicly available due to institutional and patient privacy restrictions but are available from the corresponding author on reasonable request and with permission from the participating institution and ethics committee.

## References

[1] Kidney Disease: Improving Global Outcomes (KDIGO) Diabetes Work Group. KDIGO 2022 Clinical Practice Guideline for Diabetes Management in Chronic Kidney Disease. Kidney International. 2022 Oct;102(4S):S1–S127.

[2] Kidney Disease: Improving Global Outcomes (KDIGO) CKD Work Group. KDIGO 2024 Clinical Practice Guideline for the Evaluation and Management of Chronic Kidney Disease. Kidney International. 2024 Apr;105(4S):S117–314.

[3] Palmer SC, Craig JC, Navaneethan SD, Tonelli M, Pellegrini F, Strippoli GFM. Benefits and Harms of Statin Therapy for Persons with Chronic Kidney Disease: A Systematic Review and Meta-analysis. Annals of Internal Medicine. 2012 Aug;157(4):263–75.

[4] Cholesterol Treatment Trialists’ (CTT) Collaboration, Herrington WG, Emberson J, Mihaylova B, Blackwell L, Reith C, et al. Impact of renal function on the effects of LDL cholesterol lowering with statin-based regimens: a meta-analysis of individual participant data from 28 randomised trials. The Lancet Diabetes & Endocrinology. 2016 Oct;4(10):829–39. Epub 2016 Jul 29.

[5] Colhoun HM, Betteridge DJ, Durrington PN, Hitman GA, Neil HAW, Livingstone SJ, et al. Effects of Atorvastatin on Kidney Outcomes and Cardiovascular Disease in Patients with Diabetes: An Analysis from the Collaborative Atorvastatin Diabetes Study (CARDS). American Journal of Kidney Diseases. 2009;54(5):810–9.

[6] de Zeeuw D, Anzalone DA, Cain VA, Cressman MD, Lambers Heerspink HJ, Molitoris BA, et al. Renal Effects of Atorvastatin and Rosuvastatin in Patients with Diabetes who have Progressive Renal Disease (PLANET I): A Randomised Clinical Trial. The Lancet Diabetes & Endocrinology. 2015;3(3):181–90.

[7] Shen X, Zhang Z, Zhang X, Zhao J, Zhou X, Xu Q, et al. Efficacy of Statins in Patients with Diabetic Nephropathy: A Meta-analysis of Randomized Controlled Trials. Lipids in Health and Disease. 2016;15:179.

[8] Zhou S, Su L, Xu R, Li Y, Chen R, Cao Y, et al. Statin initiation and risk of incident kidney disease in patients with diabetes. CMAJ. 2023 May;195(21):E729–38.

[9] Dormuth CR, Hemmelgarn BR, Paterson JM, et al. Use of High Potency Statins and Rates of Admission for Acute Kidney Injury: Multicenter, Retrospective Observational Analysis of Administrative Databases. BMJ. 2013;346:f880.

[10] Suissa S. Immortal Time Bias in Pharmacoepidemiology. American Journal of Epidemiology. 2008;167(4):492–9.

[11] Ray WA. Evaluating Medication Effects Outside of Clinical Trials: New-user Designs. American Journal of Epidemiology. 2003;158(9):915–20.

[12] Luijken K, Spekreijse JJ, van Smeden M, Gardarsdottir H, Groenwold RHH. New-user and Prevalent-user Designs and the Definition of Study Time Origin in Pharmacoepidemiology: A Review of Reporting Practices. Pharmacoepidemiology and Drug Safety. 2021;30(7):960–74.

[13] Faller B, Beuscart JB, Frimat L, des néphrologues de l’Est A. Competing-risk Analysis of Death and Dialysis Initiation Among Elderly (≥ 80 years) Newly Referred to Nephrologists: A French Prospective Study. BMC Nephrology. 2013;14:103.

[14] Ayav C, Beuscart JB, Briançon S, Duhamel A, Frimat L, Kessler M. Competing Risk of Death and End-stage Renal Disease in Incident Chronic Kidney Disease (Stages 3 to 5): The EPIRAN Community-based Study. BMC Nephrology. 2016;17(1):174.

[15] Fine JP, Gray RJ. A Proportional Hazards Model for the Subdistribution of a Competing Risk. Journal of the American Statistical Association. 1999;94:496–509.

[16] Lau B, Cole SR, Gange SJ. Competing Risk Regression Models for Epidemiologic Data. American Journal of Epidemiology. 2009;170(2):244–56.

[17] Reich C, Ostropolets A, Ryan P, Rijnbeek P, Schuemie M, Davydov A, et al. OHDSI Standardized Vocabularies: a large-scale centralized reference ontology for international data harmonization. Journal of the American Medical Informatics Association. 2024 Feb;31(3):583–90.

[18] Lin YC, Lai TS, Chen YT, Chou YH, Chen YM, Hung KY, et al. Comparative efficacy and choice of lipid-lowering drugs for cardiovascular and kidney outcomes in patients with chronic kidney disease: A systematic review and network meta-analysis. Journal of the Formosan Medical Association. 2024. Online ahead of print.

[19] Fravel MA, Ernst ME, Woods RL, Orchard SG, Polkinghorne KR, Wolfe R, et al. Effects of statins on kidney function in older adults. Journal of the American Geriatrics Society. 2025 Apr;73(4):1082–93. Epub 2024 Dec 18.

[20] Shin JI, Fine DM, Sang Y, Surapaneni A, Dunning SC, Inker LA, et al. Association of Rosuvastatin Use with Risk of Hematuria and Proteinuria. Journal of the American Society of Nephrology. 2022 Sep;33(9):1767-77. Article category: Clinical Epidemiology: Pharmacology and Therapeutic Development.

[21] Ward FL, John R, Bargman JM, McQuillan RF. Renal Tubular Toxicity Associated With Rosuvastatin Therapy. American Journal of Kidney Diseases. 2017 Mar;69(3):473–6. Epub 2016 Nov 14.

